# Ambulatory Video EEG extended to 10 days: A retrospective review of a large database of ictal events

**DOI:** 10.1101/2023.04.12.23288496

**Authors:** Victoria Wong, Timothy Hannon, Kiran M. Fernandes, Dean R. Freestone, Mark J. Cook, Ewan S. Nurse

**Author notes:** Contact Prof. Mark Cook, Address: 404, Clinical Sciences Block, St Vincent’s Hospital Melbourne, 41 Victoria Parade, Fitzroy VIC 3065, Australia.

## Abstract

**Objective:** This work aims to determine the AVEM duration and number of captured seizures required to resolve different clinical questions, using a retrospective review of ictal recordings.

**Methods:** Patients who underwent home-based AVEM had event data analyzed retrospectively. Studies were grouped by clinical indication: seizure differential diagnosis, classification, or treatment assessment. The proportion of studies where the conclusion was changed after the first seizure was determined, as was the AVEM duration needed for at least 99% of studies to reach a diagnostic conclusion.

**Results:** The referring clinical question was not answered entirely by the first event in 29.56% (n=227) of studies. Diagnostic and classification indications required a minimum of 7 days for at least 99% of studies to be answered, whilst treatment-assessment required at least 6 days.

**Conclusions:** At least 7 days of monitoring, and potentially multiple events, are required to adequately answer these clinical questions in at least 99% of patients. The widely applied 72 hours or single event recording cut-offs may be insufficient to correctly answer these three indications in a substantial proportion of patients.

**Significance:** Extended duration of monitoring and capturing multiple events should be considered when attempting to capture seizures on AVEM.

## Introduction

Ambulatory Video-EEG monitoring (AVEM) is an important component of epilepsy differential diagnosis, classification, and management. According to the International League Against Epilepsy (ILAE), the diagnosis of epilepsy is a clinical decision – one that can be achieved with careful history-taking and the analysis of interictal epileptiform discharges (IEDs), with investigative tests secondary (Gaillard et al., 2011). However, where epilepsy diagnosis or classification proves challenging, current ILAE guidelines recommend the use of long-term video-EEG monitoring (VEM) (Tatum et al., 2022). Compared with a routine EEG, long-term AVEM benefits with a higher likelihood of capturing seizures, moreover capturing them in a “natural” setting. The ambulatory setting often allows more appropriate assessment of events, as in-patient medication changes, removal from one’s habitual environment, and the potential stresses of inpatient provocative procedures such as sleep deprivation, may mask the true frequency and characteristics of one’s habitual seizures, interfering with diagnostic representation and subsequently management (Hasan and Tatum, 2021). In comparison to inpatient EEG, AVEM is more cost-effective, can result in higher yields, and can provide sleep samples, which are crucial as sleep is a potent activator of epileptiform discharges (Dash et al., 2012; El Shakankiry, 2010; Geut et al., 2017; Slater et al., 2019).

There is currently no specific guideline instructing the optimal duration of AVEM, and wide variability exists among different epilepsy services in their recommendation (American Clinical Neurophysiology Society, 2008; Kobulashvili et al., 2016; Velis et al., 2007). VEM studies conducted in the inpatient epilepsy-monitoring unit (EMU) are often 48-120 hours (Celik et al., 2020; Eisenman et al., 2005; Todorov et al., 1994), however the generalizability of these results to the home setting is uncertain given the aforementioned reasons, and the use of activation procedures which potentially reduce seizure latency time (Craciun et al., 2015; Schulze-Bonhage et al., 2022). Furthermore, clinical monitoring conducted at the in-home setting is often capped at 72 hours (Cho et al., 2019; Fox et al., 2019), leading to results potentially biased for shorter monitoring time and a convention adopted in practice for recommending 72 hours. Existing research into AVEM duration often investigates the average time until the first epileptic seizure as a proxy for quoting sufficient monitoring time (Celik et al., 2020; Foong and Seneviratne, 2016; Fox et al., 2019; Klein et al., 2021; Todorov et al., 1994). However, “time to first event” may lead to erroneous conclusions, as this may not represent the clinical episodes in question. Given the significant 30% epilepsy misdiagnosis rate, capture of more than one event is required to provide greater diagnostic certainty, especially as misdiagnosis can have adverse consequences for patients (Benbadis, 2007; Benbadis and Lin, 2008; Benbadis and Tatum, 2003; Ferrie, 2006; LaFrance and Benbadis, 2006).

Hence, it is paramount to define a duration of AVEM that can adequately answer a specific clinical indication. Using a retrospective database of ictal recordings, this study aimed to determine the duration of home AVEM studies needed to resolve the three main clinical questions for which the cohort was referred: for differential diagnosis between epileptic and nonepileptic seizures, classification of seizures, and the assessment of treatment. We assessed the percentage of studies in which the conclusion to the referral question was not made on the first event, to assess the appropriateness of time to the first event as a study endpoint. We then measured the optimal duration at which the majority (≥99%) of referrals are answered, hypothesizing that the optimal AVEM duration will depend on the specific clinical question. This retrospective analysis will assist in forming time-effective clinical recommendations for conducting home AVEM studies in the future, as well as reducing the delay in time that patients may experience before accessing appropriate care.

## Methods

### 2.1 Study design

This retrospective study was conducted using a home-based AVEM service (Seer Medical Pty Ltd). Data included AVEM recordings from 3,407 patients undertaken between April 2020 – April 2022 across Australia. Patients (aged 2 – 92 years, median age 34 yrs, 60.3% female) included were referred with a specific indication to the diagnostic service that met reimbursement criteria through the Australian Medicare system. Eligibility criteria included patients with:

○ A history of multiple events, and,
○ A previous routine EEG having been conducted

### 2.2 Data collection

Referrals from tertiary centers, private neurologists, pediatricians, and pediatric neurologists were made to a commercial ambulatory monitoring service (Seer Medical Pty Ltd). The AVEM technology utilized by all patients included: a wearable, ambulatory ECG/EEG monitor, a single remote camera monitor, and a hub from which data was streamed to a cloud-based ECG/EEG data repository (Nurse et al., 2023). A mobile app was provided to enable patients to report and annotate events or was provided through a paper diary depending on patient preference. Activating procedures were not used, and medication doses were not altered. After the monitoring period of 1-10 days, the data was reviewed by neurophysiology scientists, interpreted by a board-certified neurologist, and a conclusive report was created. Events were either reported by patients during AVEM or discovered by clinical review. Automated software using machine learning technology to detect epileptiform transients and seizures were employed to assist data review (Clarke et al., 2021; Eden et al., 2020).

The clinical indication for the study and AVEM data (event time-points, descriptive EEG correlate, and neurologist conclusion) was collected from the database. This data was then de-identified and recorded into a Microsoft Excel database for analysis.

### 2.3 Data analysis

Patients were first separated into epileptic or non-epileptic categories, based on the corresponding neurologist report. For the purposes of this work, only data from patients with one or more epileptic seizures were included (n= 1,653, 48.5%). Patients with diagnostic findings based purely on interictal findings (n=931) were then excluded, as this study was focused on conclusions with ictal findings. Patients with AVEM indications besides diagnostic, characterization, or treatment-assessment, were excluded (n=14), leaving 768 records labelled with their respective clinical indicators.

The three clinical indications were selected because they represent the current Australian Medicare indications for AVEM, and were the specific indications for which patients were mainly referred for and accepted for, at the home AVEM service (Item 11005 | Medicare Benefits Schedule, n.d.). Given the retrospective nature of the study and cohort eligibility being contingent on fulfilling this referral criteria, these three indications were used as the study end-points, although we acknowledge there are various alternative indications for AVEM. The final event that resolved the corresponding clinical indication, as specified in the neurologist’s conclusion, was then identified for every patient and the time-point (hour) at which this occurred was recorded. It was possible for patients to have two clinical indications (e.g. both characterization and treatment assessment), and when this was the case (n=60), the two corresponding time-points were determined for the one patient, and independently grouped under the appropriate indication category

**“**Diagnostic” indications were interpreted as being equivalent to the ILAE AVEM indication of “differential diagnosis” (Tatum et al., 2022). This was answered at the AVEM time in which the first epileptic seizure that represented the patient’s habitual event was observed. “Characterization” indications were interpreted as the ILAE definition of the “classification” of seizures and epilepsy syndromes, that is, the time until the first seizure that had defining EEG characteristics that could identify a focal or generalized epilepsy, that also matched the final neurologist conclusion.^2^ For the purposes of this work, combined or unknown epilepsy types were not considered, and for the sake of feasibility, no further subclassifications were made beyond “focal” vs “generalized” characterizations, nor time needed to further distinguish between the types of generalized epilepsies. “Treatment assessment” was answered by the time until the first epileptic seizure in individuals with previously diagnosed epilepsy currently on ASMs, as this would indicate a treatment-refractory response. The maximum duration of AVEM provided in this study (10 days) implies this indication can only be used to falsify a claim that a patient is seizure-free, and not the converse, as complete treatment evaluation requires a longer period of recording (Duun-Henriksen et al., 2020). Figure 1 summarizes this process.

**Figure 1.**
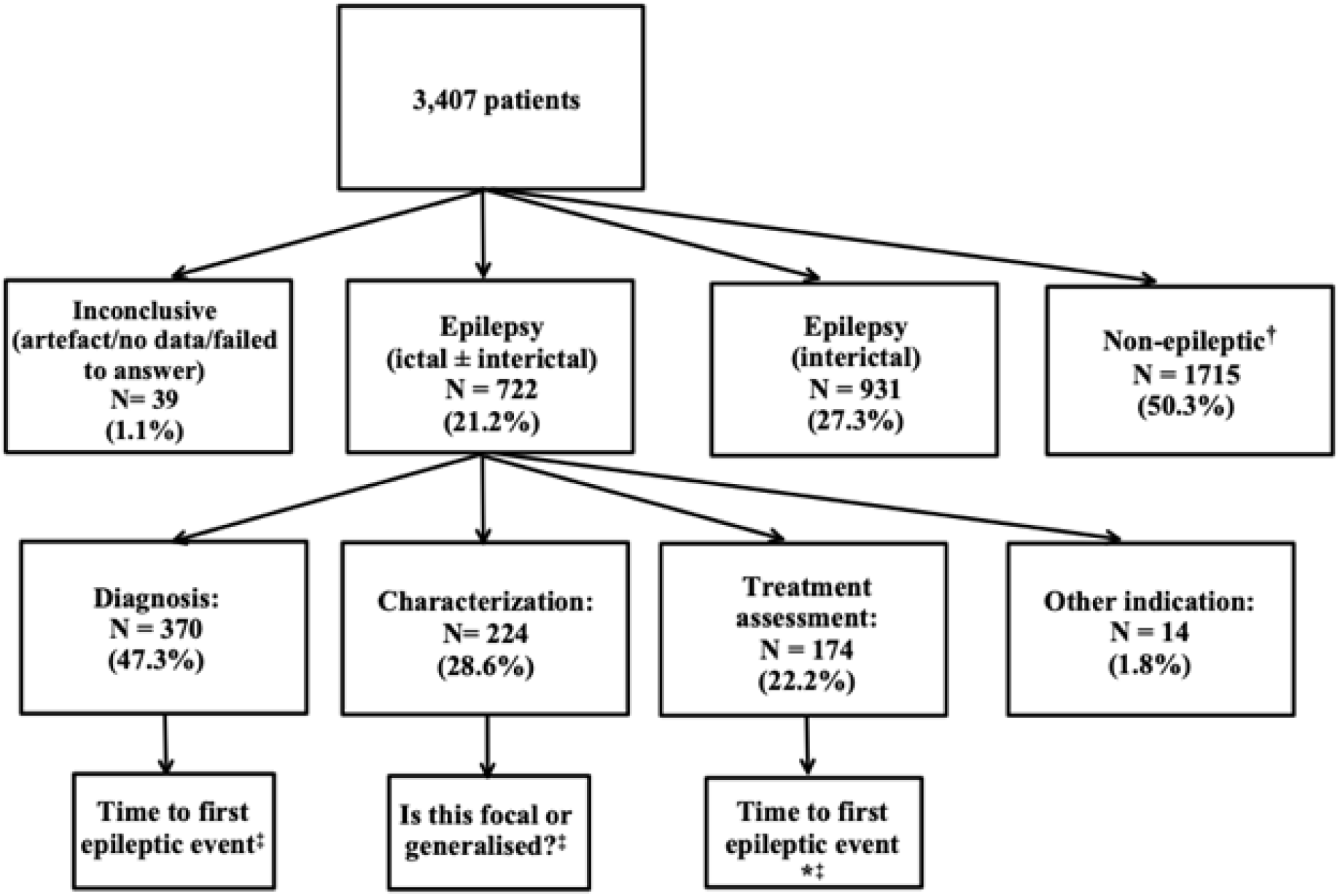
Diagram of retrospective categorization of 3,407 patients referred for AVEM between April 2020 - April 2022. Patients that had dual indications (n=60) had two independent time points identified each, and recorded into the appropriate referral group. **\***Time to first epileptic event, in patients with diagnosed epilepsy and on/previously on ASM **†**Studies that did not capture at least one epileptic event in their entirety **‡**The three primary endpoints measured in this study.

The percentage of studies answered by events occurring after the first epileptic seizure was also calculated for each group (diagnostic, characterization, treatment-assessment). These were further assessed to determine the reason behind requiring subsequent events. Categorical variables such as clinical indication and EEG characteristics were summarized using descriptive statistics like frequencies and percentages. Continuous variables such as AVEM duration were described using the median, quartiles (Q_1_, Q_3_, IQR), mean and standard deviation (SD). The average AVEM duration needed to answer each clinical indication was calculated, and given the skewed distribution of the data, the median was used to represent this. The total time needed for the majority of studies to be answered was calculated for each group, with “majority” defined as greater than or equal to 99% of studies.

Exploratory analysis was conducted to assess for significant differences in AVEM durations between the three clinical indications. The Wilcoxon Rank-sum test was chosen, as all assumptions for this test were satisfied, with the dependent variable (time duration) being continuous and not of a normal distribution, and the independent variable (the clinical indication) being categorical. A significant difference in average time was defined as p <0.05. The data analysis was conducted using Python (version 3.8.2), SciPy Statistical Package (version 1.7.1) and Microsoft Excel (version 16.63.1).

### 2.4 Ethics approval

Ethics approval for this study was approved by St. Vincent’s Hospital Melbourne Human Research Ethics Committee, under project 57392.

## Results

From 3,407 patients, 21,024 AVEM events were analyzed (80% reported, 20% discovered). 1,653 patients had epileptic seizures, and 931 of these were excluded because the conclusion was based purely on interictal findings. Of the remaining 722 patients with epileptic findings (21.2%), 60 patients had dual indications, resulting in 370 patients for diagnosis, 224 for characterization and 174 for the assessment of treatment. A breakdown of the clinical groups by study duration is provided in Table S1 of the supplementary material.

### 3.1 Subsequent event analysis

An analysis of the percentage of studies that were answered after the first seizure was undertaken, and revealed that 29.73% of diagnostic studies, 36.16% of characterization studies and 20.69% of treatment clinical questions were not conclusively answered by the first recorded seizure (Table 1). The reasons for the first event being insufficient to answer the clinical question included: an unclear first event or one partially marred by artifact, a non-habitual event, and differentiation between seizure types. Across all three referral categories, the major reason for finding utility for subsequent events, was recording new electrographic or behavioural or ictal findings. Most often the first event reported, despite described as a typical event by the patient, was shown by subsequent events to be atypical.

**Table 1.** Percentage of epileptic studies where the conclusion on epilepsy diagnosis, characterization or treatment-assessment, was based on an event after the first epileptic seizure. Each group is then broken down into reasons for the conclusion change.

|  | **Diagnostic group** | **Characterization group** | **Treatment group** |
| --- | --- | --- | --- |
| **Total percentage not answered by first seizure** | 29.73%<br>(n=110) | 36.16%<br>(n=81) | 20.69%<br>(n=36) |
| *Reason for conclusion change*<br>*(note percentages are group-wise)* |  |  |  |
| **Non-epileptic events captured before epileptic seizure** | 35.45%<br>(n=39) | 27.16%<br>(n=22) | 44.44%<br>(n=16) |
| **Different clinical or electrographic abnormality captured** | 59.09%<br>(n=65) | 66.67%<br>(n=54) | 50.00%<br>(n=18) |
| **Technical reasons** | 5.54%<br>(n=6) | 6.17%<br>(n=5) | 5.56%<br>(n=2) |

Figure 2 presents a histogram of the seizure on which the final conclusion was made. Nearly one third of studies were resolved on the second seizure, and approximately three quarters were resolved by the recording of a fifth seizure.

**Figure 2.**
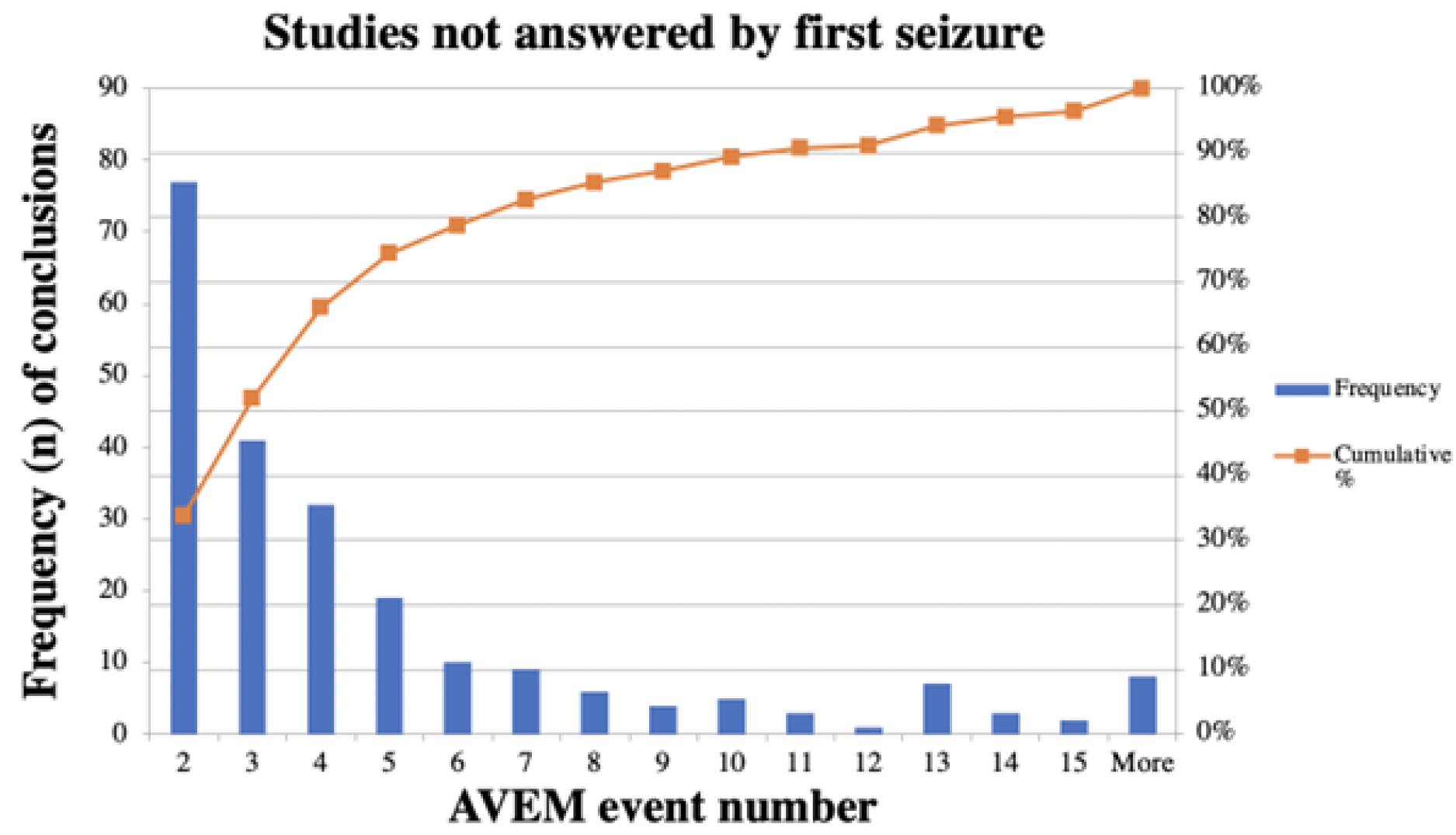
Histogram demonstrating frequency (n) of conclusions made after the first seizure. 227 studies were answered by events after the first seizure with 33.92% of conclusions being made on the second seizure. 74.45% of studies (n=169) were answered using the second to fifth seizure. 3.52% (n=8) of studies required analysis of more than 15 seizures, before a conclusion was made.

### 3.2 Diagnostic group recording duration

In the cohort of 370 patients, the median duration of AVEM needed to conclude that a patient’s typical events were epileptic was 23.25 hours (mean= 39.81hrs, SD = 40.38hrs, range = 0.01-207.15 hrs). Typically, time durations were between 9.29 hours (Q1) and 59.34 hours (Q3) (IQR =50.24hrs).

Frequency distributions (Fig 3a) display the absolute number (n) and cumulative incidence (%) of epileptic diagnoses made over each day of AVEM. The majority of diagnoses were made on day 1 (50.81%, n=188), however, a relative yield increase was associated with each additional day of recording (18.11% to day 2, 10.87% to day 3). This pattern of yield increase continued to day 4 (8.65%), after which diagnostic yield fell (5.94% to day 5, 3.25% to day 6). By day 3 (72 hrs), just under 80% of diagnoses had been made (79.73%, n=295), whilst almost all referrals (99.46%, n=368) had been sufficiently answered by the end of day 7. One patient required just over 8 days (207.15 hrs) for an epileptic seizure to be captured.

**Figure 3.**
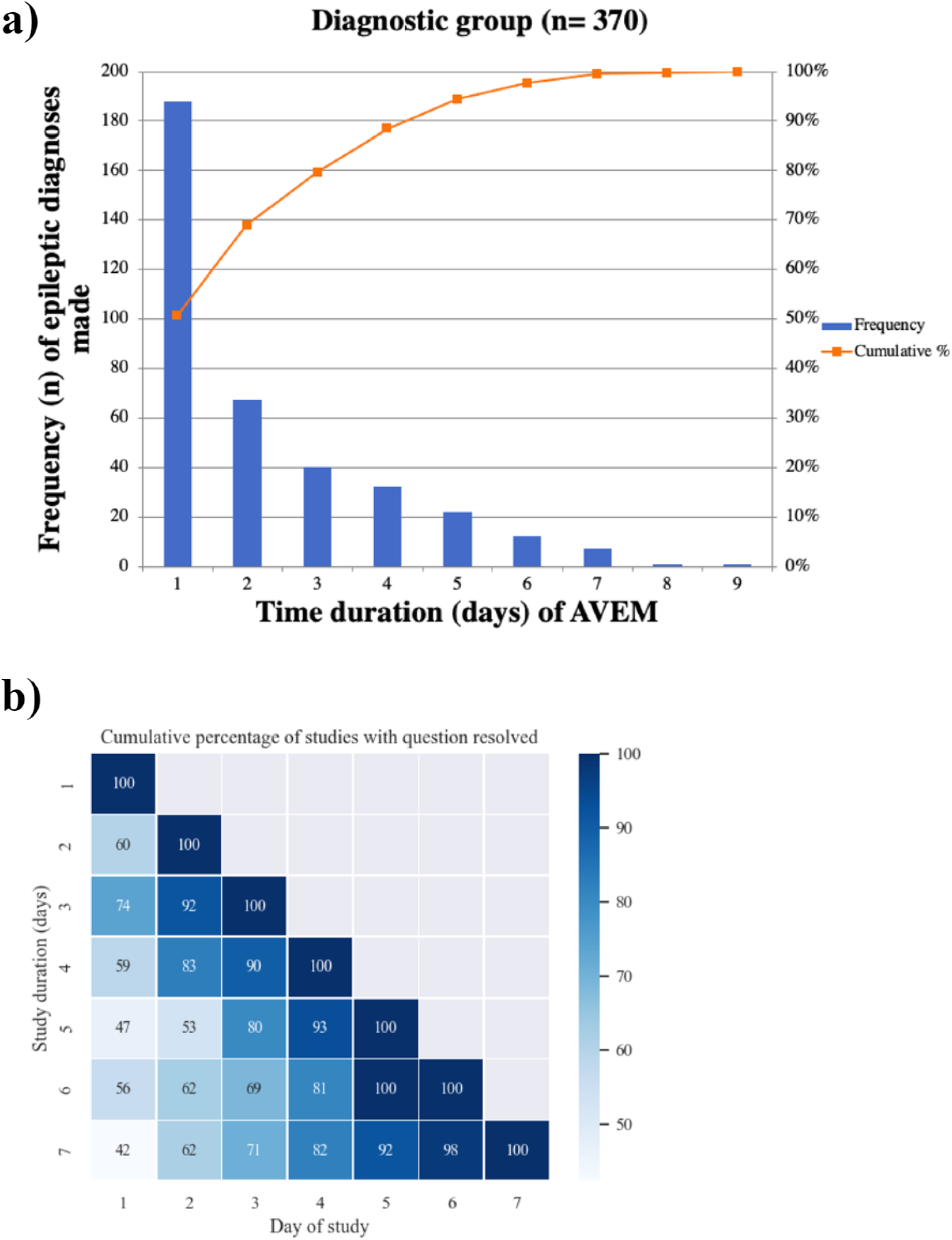
**(a)** Histogram of frequency of epileptic diagnoses made vs duration (days) of AVEM. The distribution of time durations in this sample of 370 patients is positively skewed, with a median of 23.25 hours. Note that 79.73% (295/370) of studies were answered by the end of day 3. **(b)** Heatmap of cumulative percentage of studies with diagnosis resolved. Each row corresponds to the full length of the study (days). Note that studies of more than 7 days were excluded (n=32). Further statistical information can be found in the Supplementary section.

A heatmap (Fig 3b) reveals the cumulative proportion of answered studies by day, relative to their full duration. For diagnostic studies with durations longer than 3 days (n=242, 71.6% of diagnostic studies), by day 3, 90% is the maximum proportion of studies able to be answered (90% of 4-day studies, 80% of 5-day studies, 69% of 6-day studies, 71% of 7-day studies).

### 3.3 Characterization group recording duration

Of the 224 patients indicated for further characterization of their known epilepsies, 147 (65.6%) were found to have focal epilepsies and 77 (34.4%), generalized. Median duration of AVEM needed to classify a patient’s typical seizures was 20.54 hours (mean = 39.89 hrs, SD = 42.18 hrs, range = 0.28-208.88 hrs). Typically, time durations were between 9.24 hours (Q1) and 57.97 hours (Q3) (IQR = 48.73hrs).

A frequency distribution (Fig 4a) shows day 1 had the greatest yield of epilepsy classifications completed (52.23%, n=117). Similar to the diagnostic group, there was a relative increase in the number of classifications made with each additional day after day 1 (16.97% to day 2, 11.16% to day 3, 7.59% to day 4), which fell beyond day 4 (4.46% to day 5, 3.57% to day 6, 3.57% to day 7). One patient required just over 8 days (208.88 hrs) to successfully classify their epilepsy, as no seizures had been reported or discovered before this time. By day 3, 80.36% of classifications had been made (n=180) and almost all patients had their epilepsy classification by the end of day 7 (99.55%, n=223). A heatmap (Fig 4b) shows that in characterization studies with full durations longer than 3 days (n=139, 68.81%), a maximum of 94% of studies were able to be answered sufficiently by day 3 (94% of 4-day studies, 86% of 5-day studies, 87% of 6-day studies, 69% of 7-day studies).

**Figure 4.**
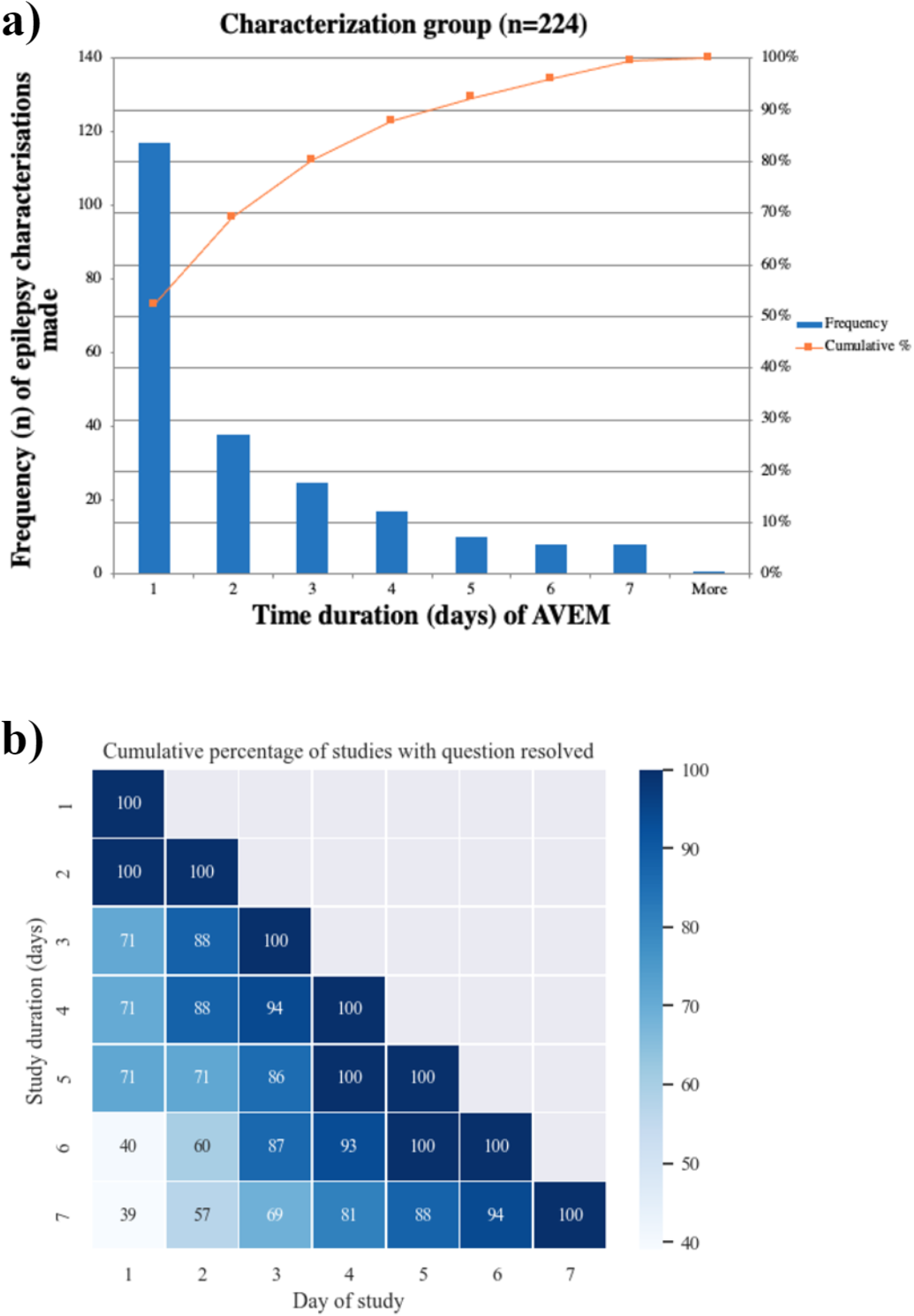
**(a)** Histogram of frequency of epileptic characterizations made vs duration (days) of AVEM. The distribution of time durations in this sample of 224 patients is positively skewed, with a median of 20.53 hours. Note that 80.36% (180/224) of studies were answered by the end of day 3. **(b)** Heatmap of cumulative percentage of studies with epilepsy characterization completed. Each row corresponds to the full length of the study (days). Note that studies of more than 7 days were excluded (n=22). Further statistical information can be found in the Supplementary section.

### 3.4 Treatment-assessment group recording duration

174 patients with previously diagnosed epilepsy were referred for AVEM to assess their response to ASM and the median monitoring time needed was 18.44 hours (mean = 30.97hrs, SD = 32.78 hrs, range = 0.28-164.56 hrs). 46.55% (n=81) of this cohort were found to have generalized seizures. Time durations were typically between 5.11 hours (Q1) and 49.11 hours (Q3) (IQR = 44.00 hrs). Of these seizures, 80.46% (n=140) were clinical seizures either identified on video review or manually reported by the patient at the time. 13.79% (n=24) of the seizures were subclinical seizures and 6.32% (n=11) were not on video or not possible to assess if clinically manifest.

Of those patients sent to confirm that seizures were controlled when none were reported, 58.62% could be shown to still be having seizures by day 1 (Figure 5a). A yield increase was associated with each additional day of recording (15.52% to day 2, 11.49% to day 3). This continued to day 4 (10.35%), after which yield decreased (1.72% to day 5). By the end of day 3 (72 hrs), 85.63% of patients had been sufficiently assessed (n=149), and by the end of day 6, almost all patients had been assessed (99.43%, n=173). A heatmap (Fig 5b) reveals by the end of day 3, 100% of 5-day duration studies were answered, compared with 91% of 4-day studies, 50% of 6-day studies and 74% of 7-day studies.

**Figure 5.**
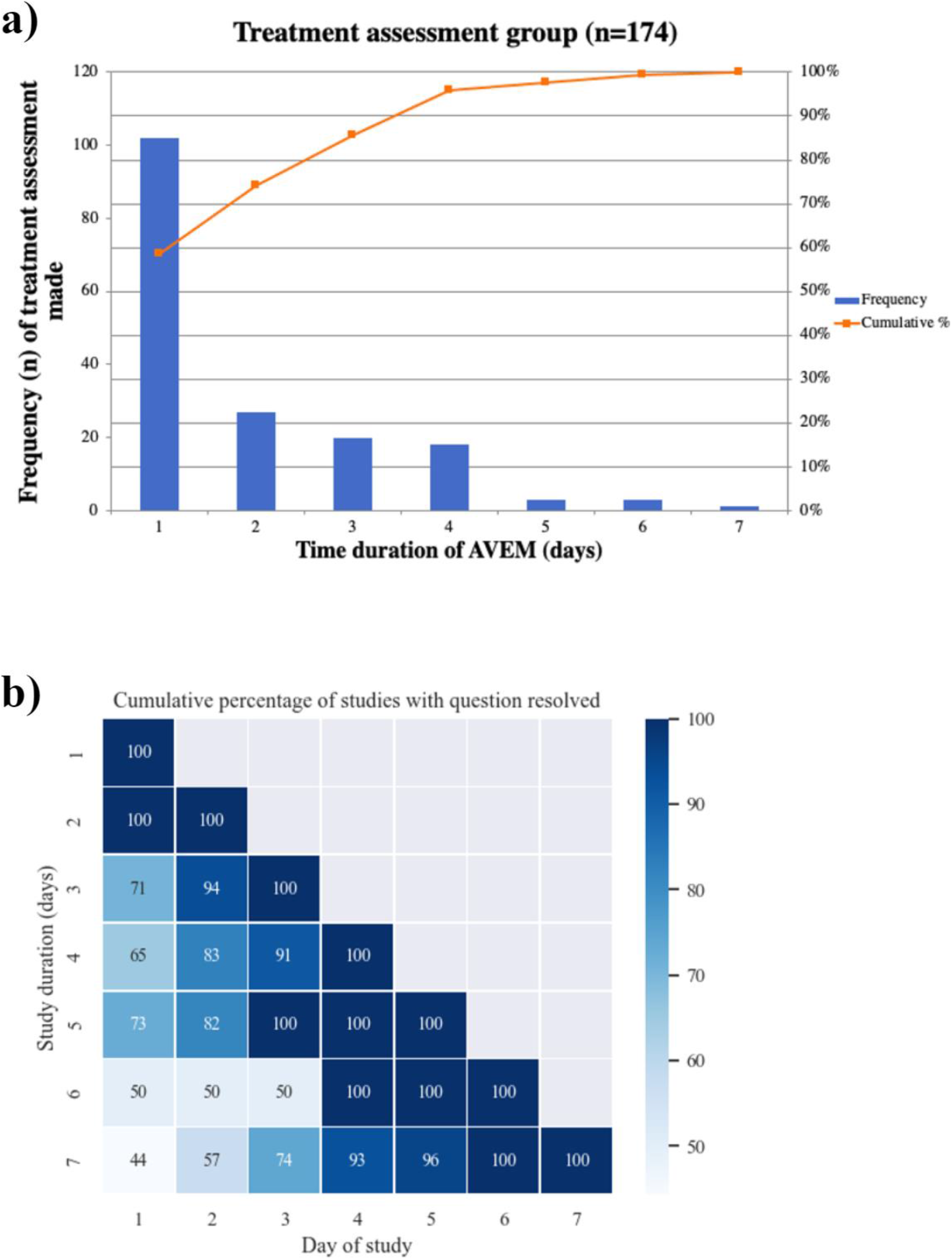
**(a)** Histogram of frequency of treatment-assessment made vs duration (day) of AVEM. The distribution of time durations in this sample of 174 patients is positively skewed, with median 18.44 hours. Note by day the end of day 3, 85.63% (149/174) of studies were answered. **(b)** Heatmap of cumulative percentage of studies that had treatment assessment completed. Each row corresponds to the full length of the study (days). Note studies of more than 7 days duration were excluded (n=10). Further statistical information can be found in the Supplementary section.

### 3.4 Recording duration difference between groups

Exploratory analysis was conducted to assess for significant differences in optimal AVEM durations between the three clinical indications, and between generalized and focal epilepsies within the characterization cohort. A 2-sided Wilcoxon rank-sum test (Fig 6) showed that the time duration needed to answer diagnostic indications varied significantly from the time needed to answer characterization questions (U= -9.215, p<0.001, 2-sided) and the time needed for treatment assessment (U = -11.041, p<0.001, 2-sided). This equated to a difference in median recording of 2.71 and 4.81 hours, respectively. AVEM durations to answer characterization questions did not differ significantly from the time needed to assess treatment (U=1.69, p=0.09, 2-sided), with an equivalent of 2.10 hours difference. Within the characterization cohort, the AVEM time needed to make a classification differed significantly in patients found to have focal epilepsies (median = 34.22hrs, n=147), compared with generalized epilepsies (median = 10.96hrs, n=77), with a median difference of 23.27 hours (U= 3.53, p<0.001, 2-sided).

**Fig 6.**
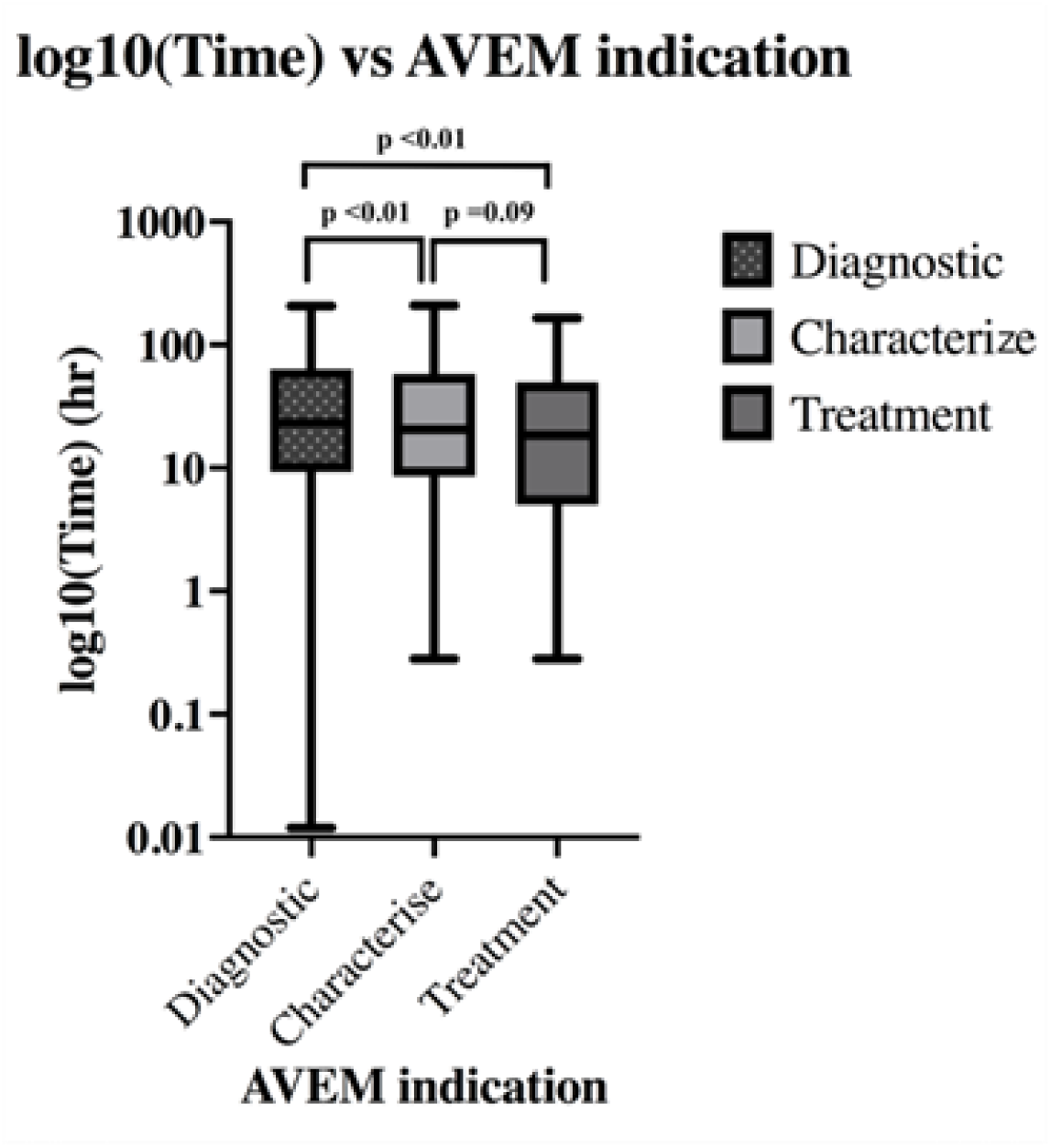
Boxplots of AVEM time to resolve clinical questions. A Wilcoxon-rank sum test was performed in this sample of 768 patients, with significance defined as p<0.05. Significant differences were found between diagnostic and characterization groups, and between diagnostic and treatment groups. Characterization and treatment groups did not differ significantly. Diagnostic indications generally took longer than non-diagnostic indications. Further statistical information can be found in the Supplementary section.

## Discussion

Our study has used a large retrospective database of ictal recordings to determine the average AVEM time needed to answer the three main indications of seizure differential diagnosis, classification, and treatment-assessment. The median durations needed to resolve all clinical questions were found to all be just under 24 hours, however, the duration at which the majority of indications (≥ 99%) is answered may be more instructive to practice. Consistent with previous studies (Foong and Seneviratne, 2016; Klein et al., 2021), after 24 hours of recording, there remained a significant proportion of patients who were yet to have an answer to their clinical question, as almost half (49.19%) of diagnoses were made after day 1. We determined that 7 days of AVEM for diagnosis and characterization, and 6 days for treatment-assessment, may be sufficient to provide definitive insight to answer the initial referral question in the vast majority of this cohort. Of course, the true “optimal” AVEM duration would depend also on the degree of completeness required by the AVEM service, as it may not be represented by a 99% coverage.

The differences in average monitoring time needed between certain clinical indications is significant, with diagnostic indications (which investigate if a habitual event is an epileptic seizure or not) needing significantly more time than non-diagnostic indications (characterization and treatment-assessment). Characterization and treatment studies included a cohort of patients with previously diagnosed epilepsy, who may have been better at recognizing, and thus, reporting their habitual events, leading to earlier seizure capture.

Treatment studies in particular often included patients who experienced seizures at higher frequency and included those being worked up for drug-resistant epilepsy. This may all contribute to shortening the length of studies where the patient has a known epilepsy compared to diagnostic studies (Mikhaeil-Demo et al., 2021; Smith, 2005), and warrants further research comparing the true reporting rates between the newly-diagnosed and the previously-diagnosed. The clinical implication is that diagnostic studies in particular may benefit from longer-term AVEM, which is feasible and favoured particularly in patients found to have epileptic seizures (Moseley et al., 2015; Nurse et al., 2023). It is noted that whilst the Wilcoxon rank-sum test has concluded significance, the true effect size may be small when translated to clinical practice (a median difference of 2.7-4.8 hrs).

There is also a significantly shorter AVEM time needed to classify epilepsies if they are generalized (median = 10.96hr, mean =21.86hrs), compared to focal (median =34.22 hrs, mean = 49.90hrs). This is consistent with research that has reported generalized epilepsies have statistically significant, shorter latencies until epileptiform discharge and require shorter total VEM durations, than focal epilepsies (Koc et al., 2019). This may be due to a clinical presentation (e.g. loss of consciousness) better recognized by both video and EEG (e.g. classic 3Hz spike-wave) as well as the increased interictal activity. Further research is warranted in exploring the impact of such factors, as well as examining the AVEM time variation between subclasses beyond simply focal and generalized. This may need to be considered when recommending optimum AVEM durations in individuals who wish to further classify their epilepsy syndromes.

Previous studies have suggested 48-72 hours of in-home AVEM is sufficient to capture typical clinical events (Klein et al., 2021; Lee et al., 2013). Such findings have contributed to a common convention in AVEM practice of “recording for 72 hours”. Our study has demonstrated that, unless AVEM is indicated in patients for ASM assessment, capping AVEM to 72 hours may lead to missed diagnoses and inadequate epilepsy classification - indications that may have been sufficiently answered should the study have extended to 7 days. AVEM research that is capped at 72 hours potentially misses conclusive data (between 14-21% of studies), which may influence the data spread and hence, median AVEM time recommended. The issue of inappropriately short recording durations is of critical importance, as misdiagnosis from inadequate AVEM duration can lead to the long-term use of inappropriate medications, medication adverse effects, excessive financial burden and social consequences. Delayed diagnosis, in certain cohorts of patients like those with multiple recurrent seizures and those with PNES, is associated with a considerably worse prognosis (Kerr et al., 2021; Parviainen et al., 2020).

When the clinical indication is epilepsy classification, greater attention to the type of suspected epilepsy, for example focal, may result in advising longer AVEM, with the knowledge that focal epilepsies require longer AVEM durations than generalized epilepsies. Existing research suggests focal epilepsies experience the greatest delay until the diagnosis is made and insufficient AVEM time may be one of the reasons why, with future research needed to corroborate this correlation (Parviainen et al., 2020). Standard, longer-term AVEM is feasible in clinical practice, with recent improvements in AVEM technology supporting stable, longer-duration recording at sufficient diagnostic quality (Nurse et al., 2022).

In comparison to the literature, the average AVEM duration reported in our study is longer and the discrepancy is likely due to the distinction between the “time until the first event”, and “time needed to answer clinical questions’’, with the former being investigated in most studies (Celik et al., 2020; Eisenman et al., 2005; Foong and Seneviratne, 2016; Fox et al., 2019; Klein et al., 2021; Todorov et al., 1994). When using AVEM for differential diagnosis, the first clinical event may not provide sufficient diagnostic information, nor represent the patient’s habitual event in question. This is evidenced by our findings that almost one-third (29.73%) of diagnostic studies relied on data from subsequent epileptic seizures. Indeed, a 2016 retrospective analysis into the reasons for prolonged length of stay (>7 days) in the EMU, found that recording insufficient habitual seizures was the leading cause, twice that of other reasons like test complications (Moseley et al., 2015).

In the case of AVEM for epilepsy classification, the first seizure alone may be insufficient when confirming focal seizures with multi-focal origins, or those exhibiting focal to bilateral tonic-clonic seizures. In our study, it was not uncommon for the neurologist to reference more than one specific event of interest before forming their conclusion, with the most common reason being the capture of different clinical or electrographic abnormalities. In comparison to the other indications, AVEM for treatment assessment may be more aligned with simply capturing the first clinical seizure, as the main question is whether the patient is still experiencing epileptic seizures with ASM use. This may be the reason for 6 days being sufficient to answer at least 99% of studies, instead of 7.

The current study has several limitations. Firstly, the retrospective nature produced a “self-selected” cohort of patients who were able to complete AVEM to a sufficient standard. The home-based AVEM service utilized is not equivalent to monitoring in a tertiary center, and patients referred were worked up for predominantly common epilepsies – the more challenging “scalp negative seizures” (12-37% of patients with frontal lobe seizures) that confound ictal data interpretation may not be well represented in this cohort (Casale et al., 2022; Ramantani et al., 2016). In the general population, the rate of inconclusive studies may be increased and the time duration may be influenced by an unequal distribution in what epilepsy syndromes patients have (Pellinen et al., 2020). A second limitation lies in the exclusion of interictal findings in our study for the sake of feasibility in locating exact time points. IEDs yields important diagnostic conclusions (Smith, 2005), and many non-epileptic and interictal studies in this database still had their respective clinical questions resolved – as such, inclusion may have predicted an earlier AVEM time. Furthermore, as stated previously, a diagnosis of epilepsy based purely on careful history-taking and the analysis of IEDs (and not ictal findings) is routinely achieved (Tatum et al., 2022). Moreover, the use of AVEM to capture IEDs before ASM initiation and discontinuation in seizure-free patients can inform management. There is much to acknowledge regarding the importance of IEDs, however seizures were the primary focus as the end-point in our study, and challenges with IED sensitivity and ill-defined criteria confound its utility in this study (Cho et al., 2019; Kural et al., 2020). Robust exploration comparing AVEM duration with and without interictal findings is warranted in the future. Another limitation is the maximum AVEM duration being 10 days in this study, potentially introducing a ceiling effect for those studies (n=39) that were inconclusive, often due to failure to catch the patient’s typical events within the finite time frame. As such, this field may benefit from longer-term studies. Furthermore, while there is no theoretical maximum limit beyond which AVEM loses yield, a study into the cost-benefit vs diagnostic yield with increasing AVEM duration will hold practical implications for both patient and healthcare provider.

Finally, the three indications of diagnosis, characterization and treatment-assessment were used as the study end-points because of the retrospective nature of the study and the fact that only patients that fit these indications under the Medicare reimbursement scheme, were accepted. There are other indications for AVEM beyond this as described by the ILAE, such as the quantification of seizure types and presurgical assessment in patients with drug-resistant epilepsy (Tatum et al., 2022). Further research into time variation between these indications, and between further epilepsy sub-classifications, would provide a more extensive guideline for time recommendations.

## Conclusion

In-home AVEM is a crucial tool for a wide range of indications in the diagnosis and management, epilepsy and is useful in its ability to capture adequate seizures to resolve clinical questions. Hence, recommendations are needed regarding an optimal duration of monitoring. This study explored AVEM duration in the context of answering specific clinical indications, revealing at least 7 days will answer 99% of all indications. Approximately one third of reports were not concluded with the recording of a single epileptic seizure.

Shortening AVEM to three days may lead to loss of valuable insight that prevents resolution of the intended clinical question. Further investigation into specific epilepsy subtypes and AVEM indications, will better solidify optimal time recommendations. This will serve to maximize efficiency in clinical evaluation, and minimize the delay before appropriate patient care is accessed.

## Data Availability

All data produced in the present study are available upon reasonable request to the authors

## Contribution Statement

VW: Data analysis and interpretation, preparation of figures and tables, manuscript preparation and editing

TH: Data analysis, manuscript editing

KMF: Data analysis, manuscript editing

DRF: Manuscript editing, data interpretation, study conception

MJC: Manuscript preparation and editing, data interpretation, study conception

ESN: Data analysis and interpretation, preparation of figures and tables, manuscript preparation and editing, study conception

