## Supplementary for "Ambulatory Video EEG extended to 10 days: A retrospective review of a large database of ictal events"

Supplementary Material

| **Duration (days)** | **Diagnostic (n)** | **Characterization (n)** | **Treatment (n)** |
|---|---|---|---|
| 1 | 6 | 4 | 6 |
| 2 | 5 | 7 | 2 |
| 3 | 85 | 52 | 48 |
| 4 | 29 | 17 | 23 |
| 5 | 15 | 7 | 11 |
| 6 | 16 | 15 | 4 |
| 7 | 182 | 100 | 70 |
| Total | 338 | 202 | 164 |

**Table S1** Number of AVEM studies (n) relative to their full study duration (days). Note this only includes studies of duration 1-7 days. Note 32 diagnostic, 22 characterization, and 10 treatment studies were not included as the duration was greater than 7 days.

| Day | Frequency (n) | Cumulative % |
|---|---|---|
| 1 | 188 | 50.81% |
| 2 | 67 | 68.92% |
| 3 | 40 | 79.73% |
| 4 | 32 | 88.38% |
| 5 | 22 | 94.32% |
| 6 | 12 | 97.57% |
| 7 | 7 | 99.46% |
| 8 | 1 | 99.73% |
| 9 | 1 | 100.00% |

**Table S2a** Distribution of time (day) since the start of AVEM, until epileptic diagnosis was made in the diagnostic cohort (n=370), with cumulative percentage answered with each additional day. Note that 99% of diagnostic studies were answered by the end of day 7.

| | |
|---|---|
| **Median (hr)** | 23.248 |
| **Q1 (hr)** | 9.293 |
| **Q3 (hr)** | 59.536 |
| **IQR (hr)** | 50.243 |
| **Mean (hr)** | 39.810 |
| **SD (hr)** | 40.381 |
| **Min (hr)** | 0.0119 |
| **Max (hr)** | 207.148 |

**Table S2b** Descriptive statistics for the AVEM duration in the diagnostic cohort (n=370)**.** The median AVEM time needed to answer diagnostic studies was 23.248 hours. 50% of patients required durations between 9.293 (Q1) and 59.536 hours (Q3). The minimum time was 0.0119 hours and the maximum time was 207.148 hours.

| Day | Frequency (n) | Cumulative % |
|---|---|---|
| 1 | 117 | 52.23% |
| 2 | 38 | 69.20% |
| 3 | 25 | 80.36% |
| 4 | 17 | 87.95% |
| 5 | 10 | 92.41% |
| 6 | 8 | 95.98% |
| 7 | 8 | 99.55% |
| 8 | 0 | 99.55% |
| 9 | 1 | 100.00% |

**Table S3a** Distribution of time (day) since the start of AVEM, until epileptic characterization was made in the characterization cohort (n=224), with cumulative percentage answered with each additional day. Note that 99% of characterizing studies were answered by the end of day 7.

| | |
|---|---|
| **Median (hr)** | 20.535 |
| **Q1 (hr)** | 9.237 |
| **Q3 (hr)** | 57.965 |
| **IQR (hr)** | 48.728 |
| **Mean (hr)** | 39.890 |
| **SD (hr)** | 42.180 |
| **Min (hr)** | 0.281 |
| **Max (hr)** | 208.880 |

**Table S3b** Descriptive statistics for the AVEM duration in the characterization cohort (n=224)**.** The median AVEM time needed to answer characterization studies was 20.53 hours. 50% of patients required durations between 9.24 (Q1) and 57.97 hours (Q3). The minimum time was 0.281 hours and the maximum time was 208.880 hours.

| Day | Frequency (n) | Cumulative % |
|---|---|---|
| 1 | 102 | 58.62% |
| 2 | 27 | 74.14% |
| 3 | 20 | 85.63% |
| 4 | 18 | 95.98% |
| 5 | 3 | 97.70% |
| 6 | 3 | 99.43% |
| 7 | 1 | 100.00% |

**Table S4a** Distribution of time (day) since the start of AVEM in the treatment cohort (n=174), until sufficient post-treatment assessment is made, with cumulative percentages listed for each additional day. Note that 99% of characterizing studies were answered by the end of day 6.

| | |
|---|---|
| **Median (hr)** | 18.4369 |
| **Q1 (hr)** | 5.1077 |
| **Q3 (hr)** | 49.1109 |
| **IQR** | 44.0031 |
| **Mean (hr)** | 30.9668 |
| **SD (hr)** | 32.7832 |
| **Min (hr)** | 0.2812 |
| **Max (hr)** | 164.5578 |

**Table S4b** Descriptive statistics for the AVEM duration in the treatment cohort (n=174)**.** The median AVEM time needed to answer characterization studies was 18.437 hours. 50% of patients required durations between 5.108 (Q1) and 49.111 hours (Q3). The minimum time was 0.281 hours and the maximum time was 164.558 hours.

| Group 1 | Group 2 | Median difference (hr) | U-statistic | Significance (p) |
|---|---|---|---|---|
| **Diagnostic** | **Characterization** | 2.713 | -9.215 | 3.10E-20 |
| **Diagnostic** | **Treatment** | 4.811 | -11.041 | 2.41E-28 |
| **Characterization** | **Treatment** | 2.098 | 1.695 | 9.00E-02 |

**Table S5a** Significant differences were found between diagnostic and characterization groups ($\mu_d$ = 2.713, U=-9.215, $n_1$ = 370, $n_2$ =224, Wilcoxin Rank-Sum test, p<0.01), and between diagnostic and treatment groups ($\mu_d$ =4.811, U=-11.041 , $n_1$ = 370, $n_2$ =174, p<0.01). Characterization and treatment groups did not differ significantly ($\mu_d$ =2.098 , U=1.695 , $n_1$ = 224, $n_2$ =174, p=0.09).
$\mu_d$ = difference in median, $n_1$ = sample size 1, $n_2$ = sample size 2

| Group 1 | Group 2 | Median difference (hr) | U-statistic | Significance (p) |
|---|---|---|---|---|
| **Focal** | **Generalized** | 23.266 | 3.527 | 4.21E-04 |

**Table 5b** Significant differences in AVEM duration were found in the characterization cohort (n=224), between focal (median = 34.22 hrs, n=147) and generalized groups (median = 10.96 hrs, n=77) (Wilcoxin Rank-Sum test).
